# Environmental, Socioeconomic, and Health Factors Associated with Gut Microbiome Species and Strains in Isolated Honduras Villages

**DOI:** 10.1101/2023.04.11.23288404

**Authors:** Shivkumar Vishnempet Shridhar, Francesco Beghini, Marcus Alexander, Adarsh Singh, Rigoberto Matute Juárez, Ilana L. Brito, Nicholas A. Christakis

## Abstract

Despite a growing interest in the gut microbiome of non-industrialized countries, data linking microbiome features from such settings to diverse phenotypes remains uncommon. Using metagenomic data from a community-based cohort of 1,871 people from isolated villages in the Mesoamerican highlands of western Honduras, we report novel associations between bacterial species and phenotypes. We also find an uncharacterized *Lachnospiraceae* species associated with 22 different phenotypes, and little overlap with a prior Dutch Microbiome Project. Furthermore, including strain-phylogenetic information modifies the overall relationship between the gut microbiome and the phenotypes, especially in some phenotypes like household wealth. Coincidentally, wealthier individuals also have a higher number of polymorphic sites. Our analysis suggests new roles that gut microbiome surveillance can play in understanding broad features of individual and public health.

## INTRODUCTION

Thanks to long-run investments in gut microbiome research in industrialized countries, the pervasive role that the human microbiome plays in influencing health-related and other phenotypes, or how, reciprocally, various phenotypes may influence the microbiome, is becoming increasingly clear (*1*,*2*). However, the majority of the human population lives outside of North America and Europe, and nearly half of the human population lives outside urban areas. Non-industrialized populations often experience problems with access to healthcare resources, have distinctive patterns of social interactions (e.g., low population density, fewer contacts with strangers), and have other distinctive exposures (e.g., animals and diet) (*3–5*). Prior studies of non-industrialized populations have documented the presence of rich uncharacterized taxa that are often absent in industrialized cohorts (*6*). And advances in genomics (such as strain-level methods) are still uncommonly applied to cohorts from non-industrialized settings. Here, we elucidate the relationship between the microbiome, characterized at the species and strain levels, on the one hand, with a very broad array of biological and social phenotypes, on the other. And we explore the role that strain-level information may play in these relationships.

## RESULTS

### Isolated Setting in Western Honduras

The village communities in the western highlands of Honduras are geographically remote (**Figure 1A**), consisting in a large proportion of the descendants of Mayan peoples who today still form traditional face-to-face social networks and who depend on subsistence agriculture and coffee cultivation. We collected population-level data in these small communities, including deep sequencing data and a comprehensive set of both individual and community-level characteristics regarding diverse socioeconomic, psychological, and health phenotypes. Our cohort consists of 1,871 people living in 19 villages which are part of a larger cohort developed for a different original purpose (*7*). The adult population in our 19 villages ranges from 66 to 432 individuals. The average age of participants was 41 (SD=17; range: 15 - 93); 63.7% were women; and 41.8% were married. Each of our 19 villages has its own intricately connected social networks with minimal inter-village contact, and they are not only separated by distance, but also by elevation (**Figure 1A**).

**Figure 1:**
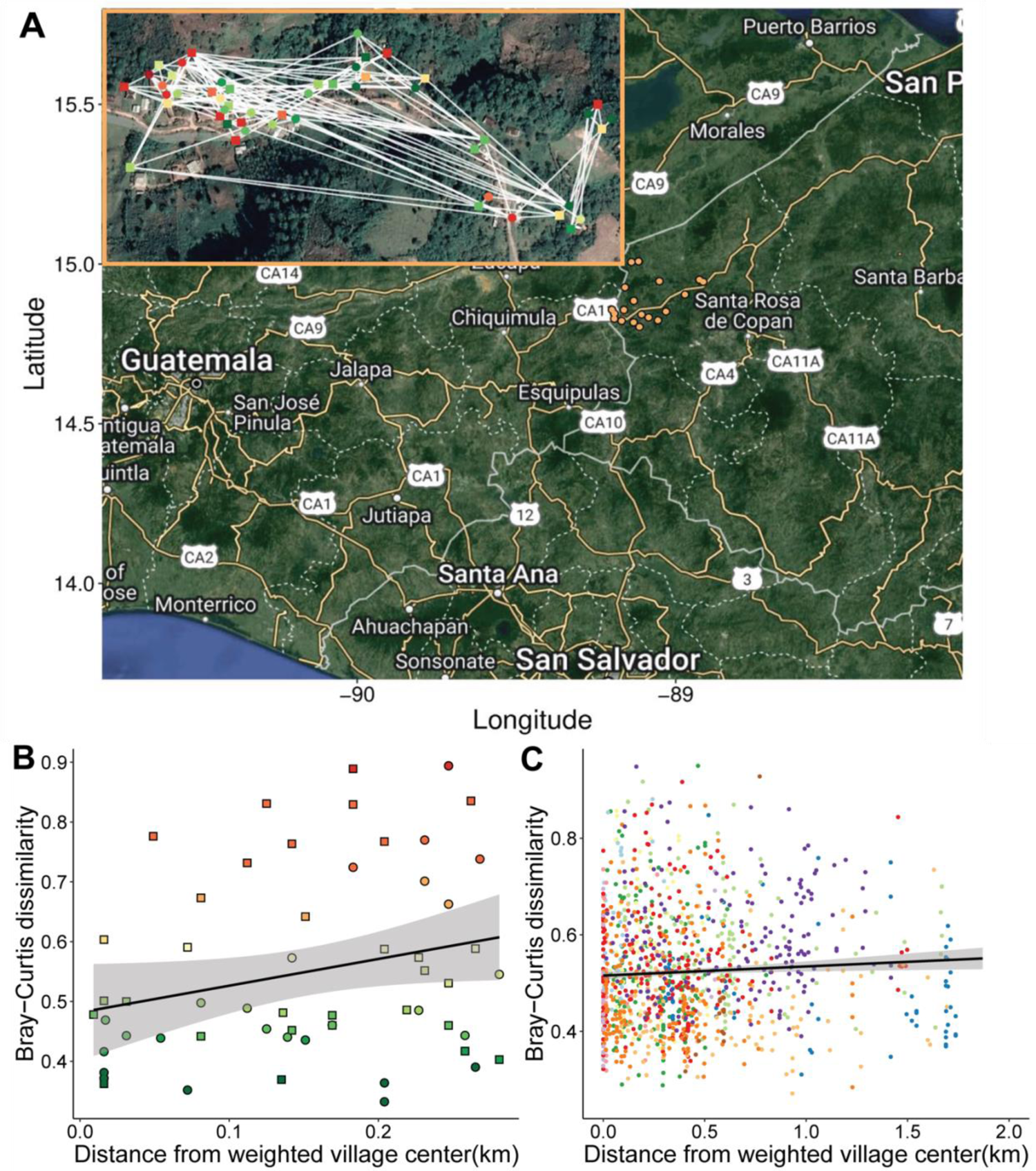
Geographic overview of the Honduras microbiome project: (A) A satellite view of the Honduran villages (in orange) that constitute the microbiome dataset. In the inset, there is a zoomed-in satellite view of an illustrative village with each inhabitant (n=57) colored with the respective Bray-Curtis dissimilarity value relative to the average microbiome composition of the rest of the village, and connected by white edges which represent social interactions between individuals. Green nodes are indicative of being very similar in microbiome composition to the rest of the village, whereas red nodes are more dissimilar. Square nodes indicate males and circle nodes indicate females. (B) Scatterplot of Bray-Curtis dissimilarity (of the village shown above) and the distance of households from the population-weighted village centroid (see Methods) shows a positive correlation (Pearson correlation coefficient ρ = 0.144, P = 0.05) between gut microbiome dissimilarity and distance from the village center. Individual dots are colored according to the person’s dissimilarity from the village’s average microbiome.

Variations in microbiome composition can be appreciated even within the same village. For instance, there is a pattern of decreasing similarity as individuals live farther away from the village center, in the geographic periphery (Pearson correlation coefficient ρ = 0.311, P = 0.0022, **Figure 1B-C**). In contrast, villagers located at the topological center of the social network within each village have a more similar microbiome to the rest of the village, unlike those at the social periphery (Linear regression β = 3.66 x 10^-05^, P = 0.761); (see Methods for details, and also the inset in **Figure 1A**).

The light grey areas indicate a 95% confidence interval. (C) Combined plot of all the Bray-Curtis dissimilarities and distances from village centroids in all the villages. The black regression line indicates a consistent trend (Pearson correlation coefficient ρ = 0.311, P = 2.2 x 10^-03^) of increasing microbiome dissimilarity with regard to the distance from the village centroid. The individual dots are colored according to the village they belong to. The light grey areas indicate a 95% confidence interval.

### Species and phenotypes

Overall, we found 2,148 associations when examining 639 microbial species and 123 phenotypes (including physical and mental health, medication use, diet, animal exposure, and social and economic measures) (**See Supplementary Table 1**). All comparisons involved appropriate statistical controls (including for batch effects; see Methods) and were corrected for multiple hypothesis testing using a False Discovery Rate (FDR) procedure. Distinctly, we also found 988 associations with metabolic pathways (**See Supplementary Table 2**). The 123 phenotypes are variously measured as continuous, categorical, and discrete variables (**Supplementary Tables 3-5**). As expected, several of the phenotype variables were correlated (for example, individuals with high Hemoglobin A1c strongly correlated with reporting a diagnosis of diabetes, and the household wealth index correlated with owning a TV (**Figure S1**)).

Similarly, the clustering of phenotypes based on species effect sizes (obtained from the species-phenotype association models) showed that multiple phenotypes within different categories have similar microbial signatures (**Figure S2**).

### Health phenotypes

We found a total of 402 species to be significantly significant associated with at least one health phenotype (**Supplementary Table 1 and 3**). Among the 402 significant species, 302 of them belonged to the phylum Firmicutes, making it the most associated with health phenotypes. Among all the associated species, 34.58% were identified as unknown (*8*) at several taxonomic levels. uSGB2239 (unknown at the genus level) from the *Rikenellaceae* family and *Parolsenella massiliensis* were the most frequently associated species, significantly associated with 5 health phenotypes; in particular, both were commonly associated with Body Mass Index (BMI), allergies, and intestinal illness (Figure 2A). uSGB2239 was also associated with antibiotics and dementia, while *Parolsenella massiliensis* was also associated with anti-hypertensive medication and openness (see Methods). Microbial species from the *Rikenellaceae* family have been previously found to be associated with at least one mental health disorder (Obsessive Compulsive Disorder) (*9*) and to be enriched in type-2 diabetics in a Pakistani cohort (*10*). In another study, *Rikenellaceae* was found to be significantly associated with Indian men having high blood sugar (*11*). Coincidentally, we found that BMI was significantly associated with uSGB2239 of the

**Figure 2:**
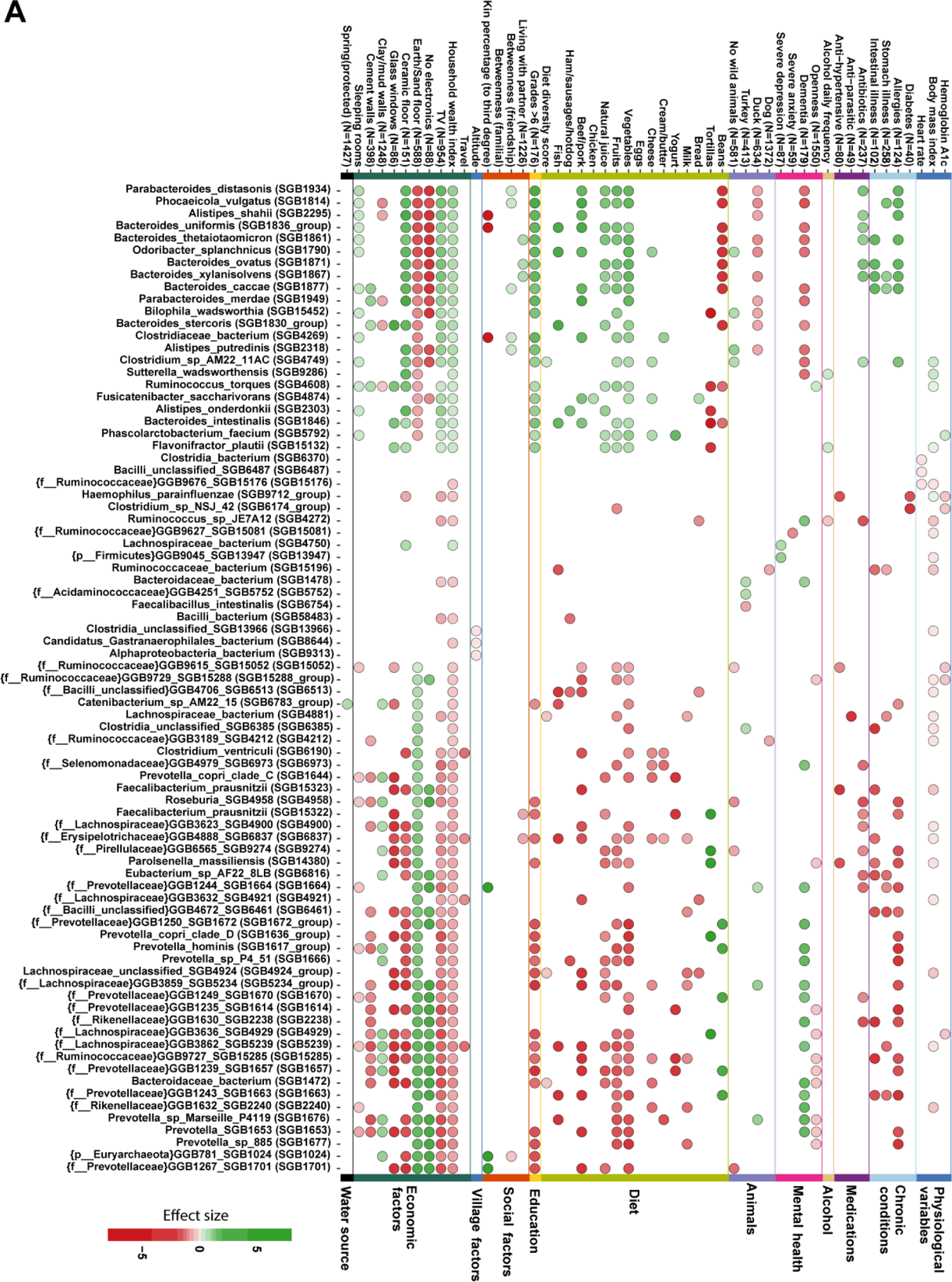
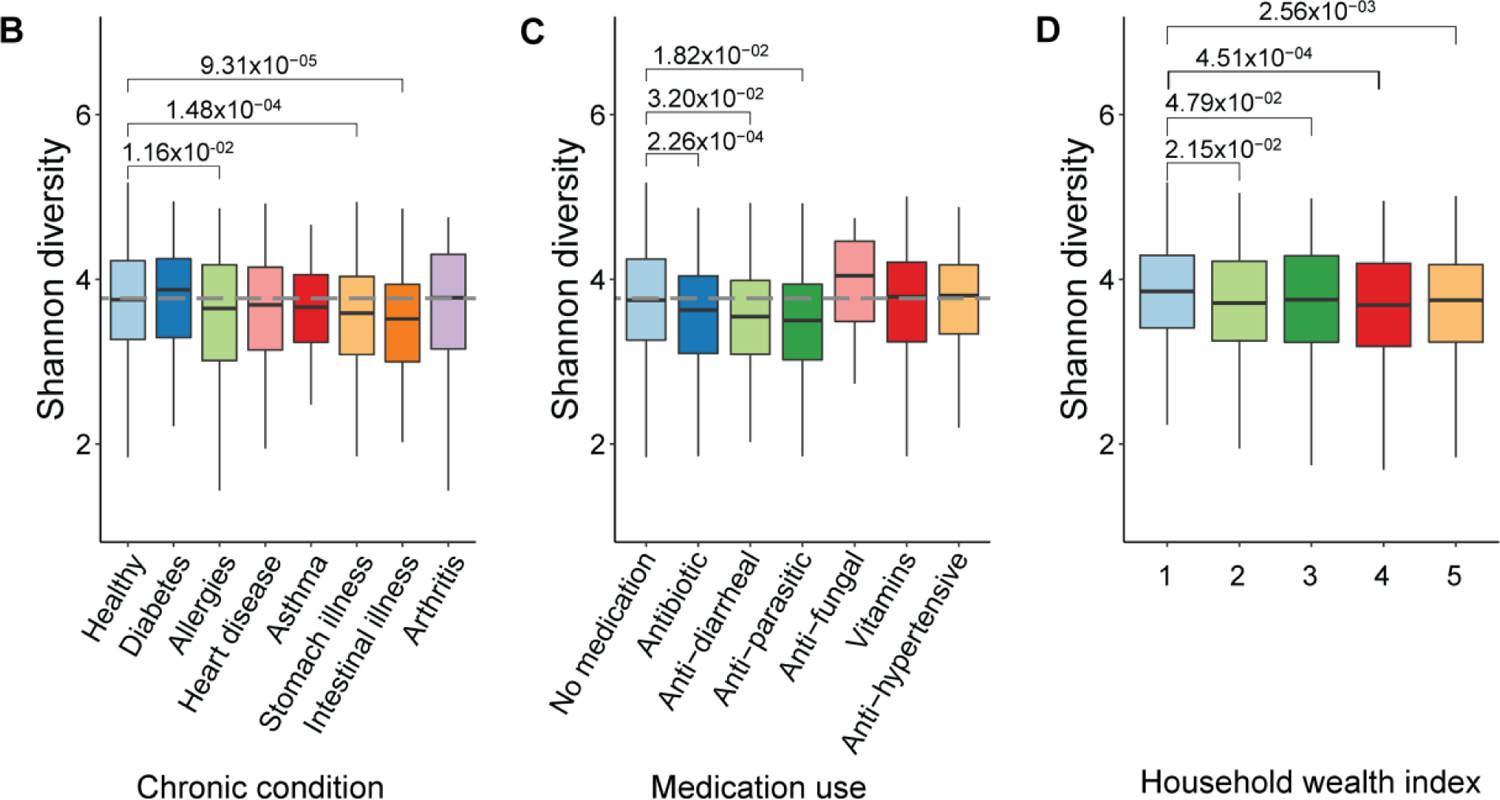
Microbiome association with phenotypes: (A) 81 species that best represent gut microbiome associations with 52 phenotypes (chosen from health, food and animal, and socioeconomic categories, see **Supplementary Table 1** for a complete list of associations). The number of individuals manifesting the respective phenotype is shown in brackets. The presence of color shows significant associations for that phenotype-species pair (FDR<0.05); the intensity of the color corresponds to the strength of the effect size. Negative associations are indicated by red color and positive by green. Unknown species are indicated with “{}”, specifying the taxonomic level at which the species is known. Phenotypes without sample size are reported for all individuals. (B) Shannon diversity of healthy and chronically ill individuals highlights differences in overall microbiome diversity; healthy individuals (n=1,407) are chosen as a reference (grey dashed line). (C) Shannon diversity is calculated between different medication use categories; non-medicated individuals (n=1,246) are chosen as reference (grey dashed line). (D) Shannon diversity of villagers belonging to households classified by household wealth index, ranging from 1 (least wealthy) to 5 (most wealthy). All comparisons were performed using the Wilcoxon Rank Sum test and corrected for multiple hypothesis testing.

### Rikenellaceae family

Furthermore, a total of 136 pathways were associated with at least one health phenotype, totaling 157 pathway associations (for details regarding the ascertainment of microbial metabolic pathways, see Methods). Among the 157 associations, physiological variables had 85 associations, followed by 24 associations in chronic illness phenotypes; 26 in medication use; 2 in acute conditions; 19 in personality measures, alcohol, cigarettes, and mental health; and 1 in overall health (**Supplementary Table 2**).

We performed association analysis for the subset of individuals falling in unhealthy ranges of various phenotypes (i.e., BMI<18 and BMI>25 to account for underweight and overweight individuals, or diastolic pressure>89 to account for hypertensive individuals) compared to healthy individuals (**Figure S3**, **Supplementary Table 6**). A total of 73 species were associated with multiple unhealthy phenotypes, of which, uSGB14313 of the Clostridia family was associated with 3 phenotypes in unhealthy ranges (Hemoglobin A1c (5.7-6.4), BMI (25–30), and BMI (30–35)) (**Figure S3A**, **Supplementary Table 6**).

We also evaluated whether the diversity of an individual’s microbiome (measured with Shannon diversity) was itself associated with various health (and other) phenotypes. The majority of the villagers self-reported themselves as healthy individuals (n=1,407, 75.20%) and only 162 villagers (8.65%) reported having more than one disease. Villagers with reported illnesses (except arthritis and diabetes) were found to have lower diversity relative to healthy villagers (Figure 2B); in particular, villagers with reported stomach (Wilcoxon Rank Sum test P = 1.48×10^-04^) and intestinal illnesses (Wilcoxon Rank Sum test p = 9.31×10^-05^) had decreased diversity. Villagers taking various medications also had lower diversity (Figure 2C); anti-parasitic drug users showed the lowest diversity (Wilcoxon Rank Sum test P = 0.018) followed by anti-diarrheal users (Wilcoxon Rank Sum test P = 0.032) and antibiotic users (Wilcoxon Rank Sum test P = 2.26×10^-04^). We found no material associations of microbiome diversity with other categories of medications.

We also performed a simple contrast analysis by comparing all chronically ill individuals to those without any chronic conditions. We used differential abundance analysis with 1,407 healthy people and 464 chronically ill people and found that 6 species were significantly differentially abundant (**Figure S3B**). *Lachnospiraceae bacterium* is the only significant species differentially enriched in healthy individuals. On the other hand, uSGB1663 and uSGB27424 of the Prevotellaceae family, *Spirochaetia bacterium*, *Coprococcus*, and uSGB6369 of the Clostridia family were found enriched in diseased individuals.

Overall, all the health phenotypes put together contribute 5.7% of the total variance explained in microbial species composition (**Figure S4, Supplementary Table 7**). Similarly, 11.6% of the variance in pathway composition is relevant to health phenotypes.

### Animal Exposure and Diet Phenotypes

We explored possible associations with animal exposure and diet (*1*, *12–14*). An unusual feature of our setting is that more than 90% of villagers reported having exposure to different types of animals, including wild animals, farm animals, and pets, affording possible zoonotic transmission. Overall, for all food and animal phenotypes, 205 species were found to be significantly associated with at least one of the phenotypes, resulting in 437 associations (Figure 2A**, Supplementary Table 4**). Among all the associating bacterial species, 27% were unknown. Among the 205 significantly associated species, 122 of them belonged to Firmicutes, making this phylum the most commonly associated with specific animals or food categories. We found 10 pathways associated with exposure to animals as well (**Supplementary Table 2**). Animal exposure contributed to 2.3% of the variation in species composition. We found no difference in overall Shannon diversity in individuals exposed to different animal categories (**Figure S5**).

Diet has been extensively studied and shown to have a substantial relationship with the gut microbiome (*14–16*). We assessed associations with microbial features and food frequency consumption and found 360 significant associations with diet (Figure 2A). *Bacteroides intestinalis* was the most associating species with food phenotypes, associating with 8 different food types. In the past, *B. intestinalis* has been implicated in the context of dietary fiber contributing to an increase of xylan utilization in the gut (*17*). Even though most of the individuals’ daily diet consist of tortillas and beans, we measured diet diversity using the Diet Diversity Score (DDS)(*18*) (see Methods and **Figure S6**). We identified a total of 108 significant associations between the DDS and gut microbiome species (Figure 2A). We also found 235 pathway associations with food phenotypes (**Supplementary Table 2**). Diet was responsible for 1.85% and 2.14% of the variance explained in our sample in species and pathways composition, respectively (**Figure S4**).

### Socioeconomic Phenotypes

Overall, we found 1,105 significant associations with socioeconomic phenotypes. For all socioeconomic phenotypes, 319 species were found to be significantly associated with at least one of the phenotypes (Figure 2A, **Supplementary Table 1**). Among all the 319 associated species, 28.8% of them were unknown and 185 of them belong to Firmicutes, making it again the most associated phylum for socioeconomic factors. Moreover, uSGB5239 of the Lachnospiraceae family is the most associated species, statistically significantly associated with 14 socioeconomic phenotypes. We also found 586 associations with pathways, with one of them associating with 9 socioeconomic phenotypes (**Supplementary Table 2**).

Socioeconomic factors are relevant to many exposures and personal habits. Higher monthly expenditures are correlated with a better diet and better household essentials such as refrigerator or a paved floor. We observed that most of the bacteria associated with higher monthly expenditures are the same as the ones associated with better diet quality (*19*, *20*).

Although all the participants in our study are considered to be living in poverty, economic status still varied among them and was associated with possessions and diets potentially relevant to the microbiome. Overall, the average household wealth index score (ranging from least wealthy (1) to most wealthy (5)) is 3.26 (standard deviation 1.33). In terms of measures of economic status, both monthly expenditure and travel were associated with the microbiome. Total wealth was also correlated with owning various items (such as a TV or a mobile phone) some of which (e.g., a refrigerator or a stove) might affect food consumption and others of which (such as having glass windows, cement walls, more sleeping rooms, an earthen floor, or a metal roof) might affect microbiome exposures via other routes (Figure 2A**, Figure S1**). We observed similar patterns of association where a high wealth index was associated with the same bacterial species associated with owning expensive items (like glass windows), and conversely. The variance explained by economic factors was 4.13% for species and 3.70% for pathways (**Figure S4**), indicating the relative importance of economic factors in explaining variation in the gut microbiome.

With respect to overall microbial diversity, the subjects from the least well-off households had a Shannon index that was increasingly higher than that of the subjects from the wealthier households (in the top 4 quintiles) (Figure 2D).

### Overall relationship between species and phenotypes

From the clustering of associations (Figure 2A) and the dendrogram (**Figure S2**), it can be observed that different phenotypes can be linked together. This link can be visualized through the relationship between the gut microbiome and phenotypes. For example, species that are enriched in socioeconomic phenotypes (such as TV ownership and household wealth index), show a similar pattern in vegetables, fruits, and meat consumption. These phenotypes also form their own collection within the dendrogram (Figure 2A, **Figure S2**); previous studies have found diet diversity to be correlated with food security and wealth in rural settings (*21*, *22*). Overall, health, food, animal, and socioeconomic factors are clustered together, which is also visibly demonstrated through the microbiome-phenotype lens. Multiple phenotypes from different categories can also be tied together by a singles species. For instance, uSGB5239 (of the *Lachnospiraceae* family) is associated with 22 different species from the health category (BMI, Stomach illness, Dementia); the food and animal category (Vegetables, Fruits, Natural juice, Beef/pork, Fish); and the socioeconomic category (Grades>6, Travel, Household wealth index, TV, No electronics, Earth/sand floor, Ceramic floor, Glass windows, Clay/mud walls, Cement walls, and Sleeping rooms) (*23*).

Furthermore, we compared our associations with the ones found by the Dutch Microbiome Project. We found 13 species-phenotype associations in common with the Dutch study, 8 of those were with BMI (Body Mass Index), and *Butyrivibrio crossotus*, *Roseburia inulinivorans*, *Faecalibacerium prausnitzii*, *Methanobrevibacter smithii*, *Eubacterium siraeum Haemophilus parainfluenzae*, *Mitsuokella multacia*, and *Flavonifractor plautii* were found significantly associated in both datasets for BMI. *Haemophilus parainfluenzae* was also significant for Hemoglobin A1c in both datasets. *Ruminococcus torques* was significant in both datasets for antibiotic use. Finally, monthly income/expenditure had 3 significant species in common: *Alistipes shahii*, *Barnesiella intestinihominis*, and *Flavonifractor plautii*. The presence of very few significant species is largely due to difference in measurements between the cohorts and the relatively low number of common species across the two cohorts (see **Supplementary Table 8 and Figure S9** for a list of possible comparisons).

### Relevance of Microbial Strains

Finally, moving beyond species-specific associations with phenotypes, we observed a meaningful variation between the genetic makeup of the same species across different individuals that is in turn associated with diverse phenotypes (Figure 3A). For instance, individuals with higher household wealth index are likely to have a different strain of *Eubacterium rectale* compared to less wealthy individuals in a set of 1,610 individuals (p=0.0004998, (Fisher’s exact test)) (Figure 3A).

**Figure 3:**
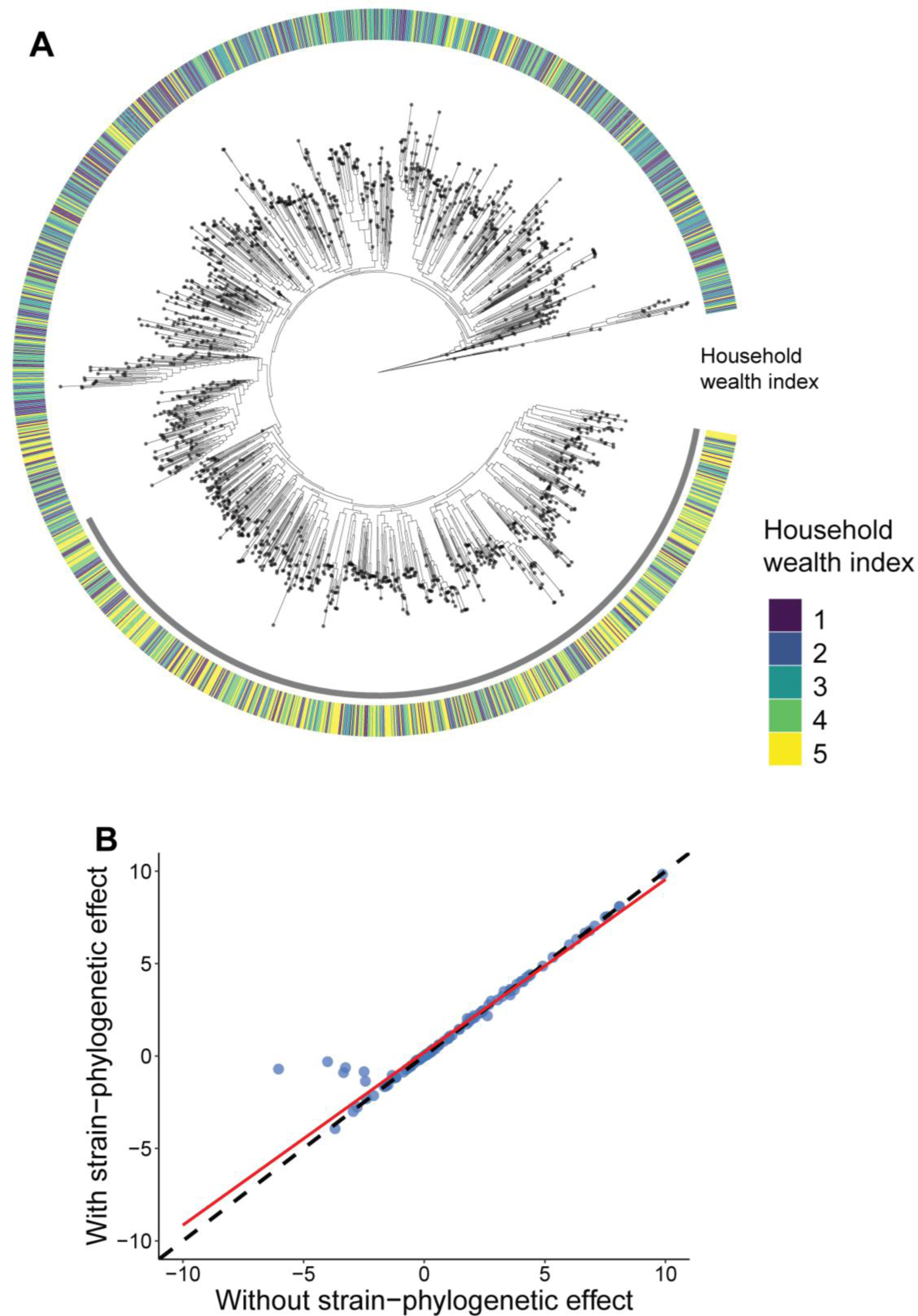
Microbial strain association with host phenotypes: (A) In this strain-level phylogeny of *Eubacterium rectale* (SGB 4933 group) (as an illustrative microbe) in 1,610 individuals, leaves are annotated with the household wealth index (as an illustrative phenotype). A cluster of individuals (annotated in grey) situated on a different strain of *Eubacterium rectale* (separated branch) are more likely to have higher wealth compared to rest of the individuals (Fisher’s exact test P=0.0004998). (B) Comparison of significant effect sizes obtained from a linear mixed model with and without adding strain-level phylogeny information across significant species-phenotype relationships overall. β-coefficients from the association models with and without including phylogenetic information are positively correlating (Spearman correlation coefficient ρ=0.989, p < 2.2×10^-16^), and the red line is the linear fit (β = 0.9351, intercept = 0.2016, p < 2.2×10^-16^), showing the relationship between the two models. The deviation of the red fitted line from the dashed line shows the important effect of adding the strain-level phylogeny in the species-phenotype association model (**Supplementary table 9**).

Moreover, adding strain-phylogenetic information in the model alters the relationship between species and phenotypes overall (Figure 3B) by inducing a small shift. Among all the effect sizes, 0.2% of them switch direction when adding the phylogenetic effect (**Figure S7, Supplementary Table 9**).

Looking deeper into the strain diversity in individuals, we looked at the variation of the percentage of polymorphic sites across individuals and phenotypes. As an illustration, we observed that individuals who are wealthier (β = 0.08345, P=1.26×10^-07^) or those consuming a higher number of eggs (β = 0.626, P=0.1146) had a higher percentage of polymorphic sites (Figure 4A-B**, Supplementary Table 10**). This comports with findings in another study where percentages of polymorphic sites from just *Prevotella copri* strains were found to be different between recent South-Asian Canadian immigrants and first-generation South-Asian Canadians (*24*).

**Figure 4:**
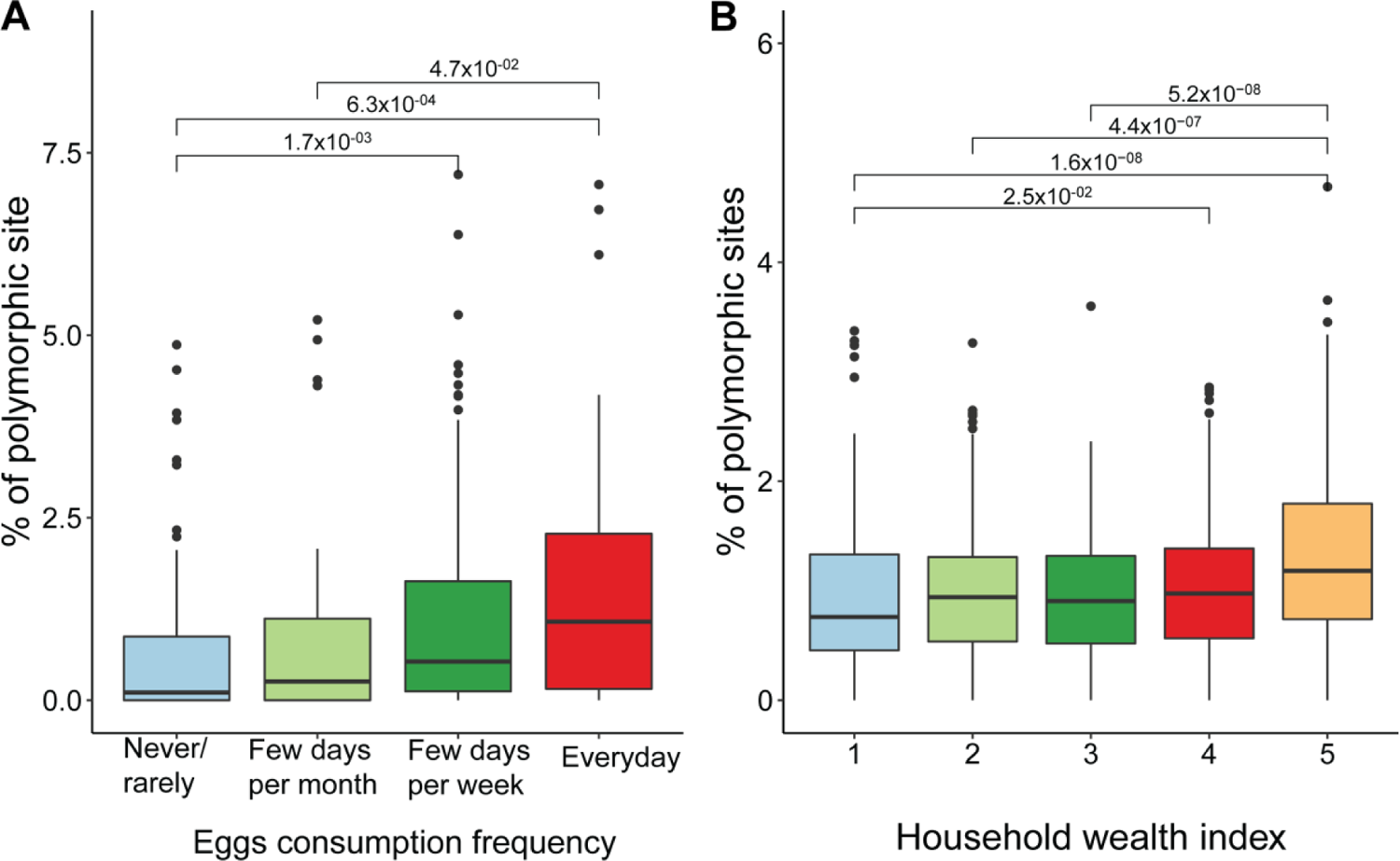
Relationship of variability in polymorphic sites with two host phenotypes: (A) Polymorphic site variability of uSGB4905 shows a gradual increase in the percentage of polymorphic sites in individuals consuming more eggs in their regular diet (N=312). (B) Another demonstration of how variability in polymorphic sites changes with phenotype. Here, progressively wealthier individuals have a higher percentage of polymorphic sites in *Prevotella copri Clade C* (SGB1644) (N=1458).

## DISCUSSION

Integrated, standardized, large, population-based cohorts to study the microbiome are uncommon, but such studies offer the prospect of identifying factors shaping the gut microbiome or being shaped by it. By extending our knowledge of the human gut microbiome to a novel population in a lower-and-middle-income (LMIC) setting; by assessing previously uncharacterized taxa; by having a very broad range of phenotypes; and by using strain-level genomic information, our goal is to advance understanding of the possible relationship of the gut microbiome with human phenotypes.

We find that variation in the gut microbiome across individuals living in a traditional way can still be at least partly explained by variations in diet, lifestyle, environment, and health factors. Overall, we found 2,148 unique associations between 639 bacterial species and 123 phenotypes. These associations included many uncharacterized species which in many cases were shown to have a stronger effect than known species. Phenotype associations were also identified after including strain-level phylogenies, which often had a profound effect on the extent of the association between microbiome species and the human phenotypes under consideration.

Still, despite measuring a large number and variety of phenotypes, only 19.2 % of the variation across individuals in microbiome composition was accounted for by these phenotypes, in keeping with prior studies (*1*, *25–27*). This suggests that microbiome composition in individuals may be quite idiosyncratic or may depend on details of social interactions or unmeasured environmental exposures. Rare species may also help account for this variation. The current understanding of how individual and population-level microbiomes come to be shaped is thus still incomplete. Nevertheless, the phenotypes we ascertained in Honduras did combine to account for 19.2% of the species variation (as noted) and 33.4% of the pathway variation; this may be compared to a recent study from the Netherlands where the measured phenotypes accounted for 13% and 16.2% of the variation, respectively, (*1*) although different methodologies for taxonomic and functional characterization were used here, reflecting ongoing methodological advances.

The gut microbiome is known to be related to various health conditions both in humans and in mice (*28*), and conditions like cancer, obesity, diabetes, autism, anxiety, and depression can induce shifts in the gut composition (as shown in many mostly Westernized populations) (*1*, *28–34*). Alcohol intake and cigarette use have been linked to gut microbiome dysbiosis, as have medications (*35–39*). In keeping with these prior studies, we confirm such findings in this rural LMIC cohort (*3–5*, *40*). Indeed, we found 606 associations between the microbiome and health-related phenotypes. Chronic illnesses and medication use were the most strongly associated. Among chronic illnesses, intestinal illnesses show the greatest differences. We uncovered 297 total associations between gut microbiome species and physiological measurement ranges that may be linked to underlying chronic conditions such as obesity, diabetes, and hypertension. Moreover, we find 64 associations with mental health phenotypes alone, a relatively understudied area.

Looking at the overall microbial composition among healthy and chronically ill subjects, the Shannon diversity was generally lower in most of the chronically ill people, especially those with allergies and gastrointestinal illnesses. Moreover, comparing healthy individuals to those who are chronically ill, we found 6 taxa to be differentially enriched in one of the groups. *Lachnospiraceae bacterium* is the only significant species differentially enriched in healthy individuals. On the other hand, 2 unknown and 3 known species: uSGB1663 and uSGB27424 of the Prevotellaceae family, *Spirochaetia* bacterium, *Coprococcus*, and uSGB6369 of the Clostridia family, are differentially enriched in chronically diseased individuals. Among medication users, those taking anti-parasitic medication had the largest drop in overall diversity.

Another factor that greatly influences the gut microbiome is diet. Our sample population exhibits a consistent diet, with beans and tortillas consumed by most people daily. Still, we found 360 associations with food categories. A previously studied Dutch cohort found that pets had notable associations with the microbiome (*1*), and we also found 77 associations with a broader range of animal exposures.

Furthermore, our sample was spread across 19 villages separated in space and elevation, and the overall gut microbiome was observed to vary with the relative spatial position within the villages; the dissimilarity score with a village-averaged microbiome increased as subjects lived further away from the village center. Relatedly, we found 3 significant associations with elevation. We also found 6 associations with other village-related factors, such as distance to the health center (which is historically correlated with overall health in middle/low-income areas) (*3*), distance to main road, and number of churches.

Social and economic factors had 1,105 associations, with the bulk of the strong associations coming from unknown species. The gut microbiome had 947 unique associations with economic factors alone, making it the second highest associating category of variables we examined, after health. Wealth differences in individuals can also manifest in the form of more diverse strains in some species being present in wealthier individuals. Prior research in Honduras has highlighted the crucial importance of socioeconomic status in addressing health in such communities (*41*), and the microbiome varies in important ways along this axis, even in this poor setting.

Social interaction is an integral part of Honduran villagers’ life. In total, 123 unique associations with various social network factors were found. Studies investigating social interactions between mice have shown the evolutionary advantage of having behavior that enhances social interaction that consequently facilitates microbiome transmission (*28*, *42*, *43*). In wild mice, social associations are predictive of microbiome composition, and the microbiome is correlated across mice interaction networks (*44*). In humans, strain-level similarities have been shown in familial and partner networks within and outside households (*45–47*). Whether these interactions translate into exposures that directly contribute to health is an important area for further work.

Uncharacterized taxa play a vital role in all these associations, as in prior LMIC cohorts (*6*). Despite the number of unknown species in the Honduran cohort being about a third of total species, their relative strength of associations was observed to be higher in all the phenotype categories. Distinctly, strain-level information is also relevant to the microbiome-phenotype relationship and should optimally be accounted for.

By expanding our knowledge of the human microbiome to a novel non-Westernized cohort, it is possible to further our understanding of the role of the gut microbiome in chronic illness and, at the same time, open up opportunities to use such findings to develop inexpensive biomarkers to aid diagnostics in rural settings (*48*, *49*). To the extent that a healthy microbiome is driven by modifiable social and environmental factors (such as diet, smoking, living arrangements, lifestyle, and so on), understanding which factors to target or what possible microbiome-modifying interventions to implement could help advance individual and collective health in diverse settings.

## METHODS

### Sample collection, library preparation, and sequencing

Participants were instructed on how to self-collect the fecal samples using a training module and promptly returned samples to a local team which then stored them in liquid nitrogen at the collection site and then moved them to a −80 C° freezer in Copan Ruinas, Honduras. Samples were then shipped on dry ice to the United States and stored in −80 C° freezers.

Stool material was homogenized using TissueLyzer from Quigen and the resulting lysate was prepared for extraction with the Chemagic Stool gDNA extraction kit (Perkin Elmer) and extracted on the Chemagic 360 Instrument (Perkin Elmer) following the manufacturer’s protocol. Sequencing libraries were prepared using the KAPA Hyper Library Preparation kit (KAPA Biosystems). Shotgun metagenomic sequencing was carried out on Illumina NovaSeq 6000. Samples not reaching the desired sequencing depth of 50Gbp were resequenced on a separate run.

Raw metagenomic reads were deduplicated using prinseq lite (version 0.20.2 (*50*)) with default parameters. The resulting reads were screened for human contamination (hg19) with BMTagger (*51*) and then quality filtered with trimmomatic (*52*)(version 0.36, parameters “ILLUMINACLIP:nextera_truseq_adapters.fasta:2:30:10:8:true SLIDINGWINDOW:4:15 LEADING:3 TRAILING:3 MINLEN:50”). This resulted in a total of 1,871 samples (with an average of 86.84 x 10^6^ reads per sample).

### Local involvement in the research

In keeping with proper standards for such research, we worked closely with the local population of Copan, sought feedback and approval from officials at the Ministry of Health (MOH) of Honduras, and endeavored to provide practical benefits to the local community. Here, we briefly summarize this history and outline some of our principles and actions in this regard.

When we began designing this cohort project in 2013 (for the whole cohort of 176 villages and 24,702 people in the parent RCT), the Bill and Melinda Gates Foundation (BMGF) introduced us to the Inter-American Development Bank (IDB), which has been supporting and doing work throughout Latin America, and IDB in turn introduced us to the Honduras MOH. After the cohort was established, we obtained additional funding from other sources as well. Because of this pathway to getting the project launched, we worked with local and regional public health organizations and with local leaders rather than with local academic institutions.

From the outset when the original underlying cohort for this study was impaneled (in 2013), we sought extensive local involvement, beginning with a needs assessment where local village residents told us about topics of concern to them in a series of meetings in villages throughout the Copan region (in the western highlands of Honduras, near the Guatemala border). In addition to extensive community input, we sought input from the MOH. We periodically briefed both the communities and the MOH about our findings.

Copan is a very isolated area, with villages within Copan being further isolated due to varying altitudes (850.13±154.77 m). Over the years, as we built our data collection team in Copan, we developed deep ties to the local community, to local village leaders, to the few local health clinics, and to local transportation and infrastructure providers. Because of these ties and our commitment to the local community, we presented our results directly to these constituencies regularly. We also held two annual joint implementation science conferences with our Honduras and Yale teams (involving many dozens of attendees).

We provided other material benefits to the local community, beyond simply providing them with information. When we tested people for parasites as part of our study, we gave them the results of their tests and arranged for them to be treated. When we tested people for vision, we provided corrective glasses. We solicited ideas from the local community about what infrastructure improvements we could make, and we repaired many local playgrounds and clinics as a result. We arranged for an American company to provide free portable handheld ultrasound devices to the local health clinics, which was much appreciated by local providers. In terms of capacity building, we hired and trained over 100 local people and built capacity in the region; and many of our former data collectors have gone on to work for other public health and development entities. Lastly, we offered a talented young person from Copan a slot as a PhD student in the USA.

Throughout our work in Honduras, and given the extent of local involvement at the regional and MOH levels, we endeavored to act with integrity, curiosity, and respect in all relationships.

Finally, we note that this research would not have been prohibited in the USA. This work is not likely to result in stigmatization, incrimination, or discrimination for the participants, and we have carefully safeguarded all data from threats to the privacy or security of our participants, which has constrained the individual-level data released here.

### Taxonomic profiling and diversity analysis

Quantification of organisms’ relative abundance was performed using MetaPhlAn 4 (*8*), which internally mapped the metagenomes against a database of ∼5.1M marker genes describing more than 27k∼ species-level genome bins (SGB).

We identified a total of 2,508 species in our dataset. Among the 2,508 species, 639 species were used for association analysis after filtering for minimum relative abundance values (10^-2^), and a minimum of 10% prevalence in the population (n=187).

We performed strain-level profiling for these species with StrainPhlAn4 (*8*)(parameters: “-phylophlan_mode accurate”)

Microbiome species richness was estimated using the Shannon entropy index and the total number of observed species (i.e., those with relative abundance simply greater than zero). Multidimensional scaling analysis (vegan cmdscale function) was performed on the Bray-Curtis dissimilarity index (vegan vegdist function) calculated on the relative abundances obtained by MetaPhlAn4.

Functional potential analysis was performed using HUMAnN 3.0. (*53*). Gene family profiles were normalized using relative abundances and collapsed into MetaCyc pathways.

To understand the amount of variance explained by various factors, we performed a PERMANOVA analysis (adonis function from the vegan package (*54*)) using the “bray” method; the diversity matrix was calculated on both species-level relative abundances and MetaCYC pathway relative abundances as input, including the 123 phenotypes variables into the model. All the comparisons were run with 999 permutations.

### Phenotype characterization

We measured a broad range of phenotypes using standard measures (*7*). Description and statistics on all phenotypes can be found in **Supplementary Tables 2-4**. Physiological measurements were deemed within normal limits in accordance with CDC (*55*) and NBME (*56*) guidelines (**Figure S3**).

We used self-reported information to discern whether people were healthy or were diagnosed with various conditions. General anxiety disorder is derived from a set of 7 questions from a self-reported survey-based questionnaire, which assigns a score of 0 to “Not at all”, 1 to “Several days”, 2 to “More than half the days”, and 3 to “Nearly every day”. The scores are added up (maximum of 21) and partitioned as: Minimal or none (≤5), Mild (6–10), Moderate (11–15), and Severe (≥16) (*57*). The PHQ9 (Patient Health Questionnaire) score measuring depression was computed similarly, with the levels being: Minimal or none (≤5), Mild (6–10), Moderate (11–15), Moderately severe (16–19), and Severe (≥20) (*58*). Personality traits like Openness or Nervousness were also based on self-reported questions, where the participants were asked to rate themselves between strongly disagree to strongly agree for each of the personality questions.

The Frequency of intake of various food items was self-reported, ranging from: “Never/rarely” to “Every day”. These frequencies were used as input in the diet-microbiome association model. The diet diversity score (DDS) (*18*) was calculated by classifying individual food types into one of the following categories: cereals, roots/tubers, vegetables, fruits, meat/poultry/offal, eggs, fish/seafood, pulses/legumes/nuts, milk and milk products, oils/fats, or sugar/honey. If any of these food items were consumed daily, the respective categories would get 1 for that individual. The sum across these categories would define the DDS score of this individual. The maximum possible DDS score would be 11 and the minimum would be 0.

Numerical values were reported for alcohol frequency and cigarette frequency. The daily alcohol intake ranged from “1 or 2” to “10 or more” drinks. Cigarette usage was reported as a “Yes” or “No”.

The household wealth index is computed using Multiple Correspondence Analysis (MCA) based on all the household items. The index ranged from 1, indicating low wealth, to 5, indicating high wealth.

We explored associations with several social network features, including the degree, transitivity, and betweenness centrality of each individual. To uncouple the effects of kin and non-kin social connections, we investigated microbiome associations in familial networks, friendship networks, and combined networks. In the combined network, we computed the amount of kin in a person’s first three degrees of social connections (i.e., among a person’s friends, friends of friends, and friends of friends of friends) to assess the relative effect of having kin close to a person within the social network. In addition to kin and non-kin relationships, we also explored the microbiome’s association with cohabiting partners.

### Population-weighted village centroid

We collected the GPS coordinates (latitude and longitudes) of all the building in the village. Since multiple individuals can reside in a building, the population-weighted centroid was chosen as the reference center of the village, which was then used to compute every individual’s distance from this village center. Satellite plots were created using “ggmap” package in R (*59*).

### Model for microbiome-phenotype regression

For the association model with species-level microbiome and phenotypes, a linear mixed model (lmerTest Rpackage (v 3.1)) was used to explore the relationship of the variability in phenotype and the variability in the microbiome.

For every species and phenotype pair, we computed the following model

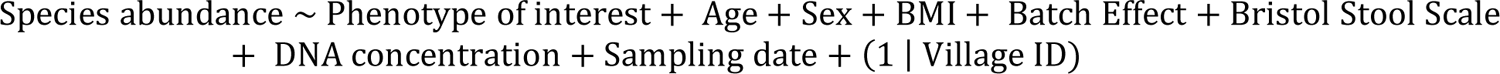

Species’ relative abundances were transformed using the CLR (Centered-Log Ratio) and used as Since basic demographic attributes (age, sex), technical factors (DNA concentration, sequencing batch, sampling date), and BMI and Bristol stool scale accounted for 1.65% of the species variation and 5.86% of the pathway variation, we used these variables as primary controls in our association models (**Figure S4**, **Figure S8**).

Furthermore, all associations were corrected for both microbiome species and phenotype using multiple hypothesis testing (Benjamini-Hochberg correction) and all significant associations are corrected for an FDR (False Discovery Rate) <0.05.

### Strain-phenotype analysis and phylogenetic signal

For strain-level analysis, we used the Almer function from the “evolvability” R package (v 2.0.0). Almer incorporates phylogenetic trees in mixed linear models as a correlated random effects structure.

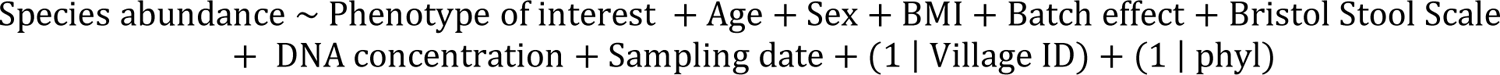

where, “phyl” is the variance-covariance matrix calculated from the species’ phylogenetic tree. To evaluate the strain-phylogenetic effect, we compared beta coefficients from this model and the same model without the random effect on the variance-covariance matrix.

The phylogenetic signal was estimated using the “phylosig” function in “phytools” R package (v 1.9-23) using the ‘lambda’ method. Overall, among the 78,597 species-phenotype pairs (639 species and 123 phenotypes), 52,864 pairs were chosen after filtering for phylogenetic signal. The phylogenetic signal was estimated for the phylogenetic tree of each species vs phenotype of interest.

### Polymorphic sites analysis

For polymorphic sites, files suffixed with “.polymorphic” in StrainPhlAn4 output were used after discarding 0’s in the “percentage of polymorphic sites” column to discard subjects without the species of interest. Wilcoxon rank-sum tests were performed across categories within phenotypes to check for significant changes in polymorphic sites. In addition, linear regression was also performed to investigate the relationship between polymorphic site percentage and individual host phenotypes (see **Supplementary Table 10**).

### Differential abundance analysis

We used the MaAsLin 2 (v 1.0.0) package in R to determine the association between species and disease status (healthy or unhealthy) of individuals and to estimate the effect sizes and adjusted p-values (FDR corrected). A list of significantly positive and negatively associating species was recorded. Species-level relative abundances were normalized (in MaAsLin2) and used as input for MaAsLin2 in which age, sex, BMI, DNA concentration, sampling date, and Bristol stool scale were used as controls and village as a random effect. All the resulting p-values obtained by the MaAsLin2 models were corrected for multiple hypothesis testing using FDR.

## Supporting information

Supplementary Informations

## Data Availability

The code for replicating the analysis is available at https://github.com/human-nature-lab/Phenotype-paper. Metagenomic sequences for the study participants are deposited in NBCI SRA and available under accession number PRJNA999635.

## Acknowledgments

We thank all the study participants in Honduras. We thank Jose Eduardo Gámez and Eduardo Jose Urrea Carbajal for coordinating the fieldwork; Rennie Negron, Liza Nicoll, and Thomas Keegan for their support on field operations, data collection, and administrative support; YCGA (Yale Center for Genomic Analysis) for sequencing the metagenomic libraries; and Qiaojuan Shi for processing the specimens and handling the extractions. We thank Michael Baym and Mark Gerstein for helpful comments on the manuscript. This work was supported by the NOMIS Foundation, with additional support from Schmidt Futures, the Pershing Square Foundation, and the Rothberg Catalyzer Fund. Empanelment of the underlying cohort was supported by the Bill and Melinda Gates Foundation.

## Author contributions

Conceptualization: SVS, FB, MA, IB, and NAC; Methodology: SVS, FB, MA, AS, IB, and NAC; Data Collection: SVS, FB, MA, RMJ, IB, and NAC; Statistical Analysis: SVS, FB, MA; Funding acquisition: NC; Supervision: IB, NAC; Writing: SVS, FB, MA, IB, and NAC.

## Competing interests

The authors declare that they have no competing interests.

## Notes

### Competing Interest Statement

The authors have declared no competing interest.

### Author Declarations

IRB of Yale University gave ethical approval for this work

### Summary of Updates

The manuscript has been revised to update the analysis method

