## Supplementary Informations for "Environmental, Socioeconomic, and Health Factors Associated with Gut Microbiome Species and Strains in Isolated Honduras Villages"

**Figure List:**

1. **Phenotype-phenotype correlation.**
2. **Phenotype-microbiome association clustering.**
3. **Relationship between health and microbiome.**
4. **Variance explained.**
5. **Alpha diversity of individuals exposed to animals.**
6. **Diet diversity score.**
7. **Comparison of species and strain models.**
8. **Principal Coordinates Analysis (PCoA).**
9. **Comparing Honduran and Dutch (Lifelines) datasets.**

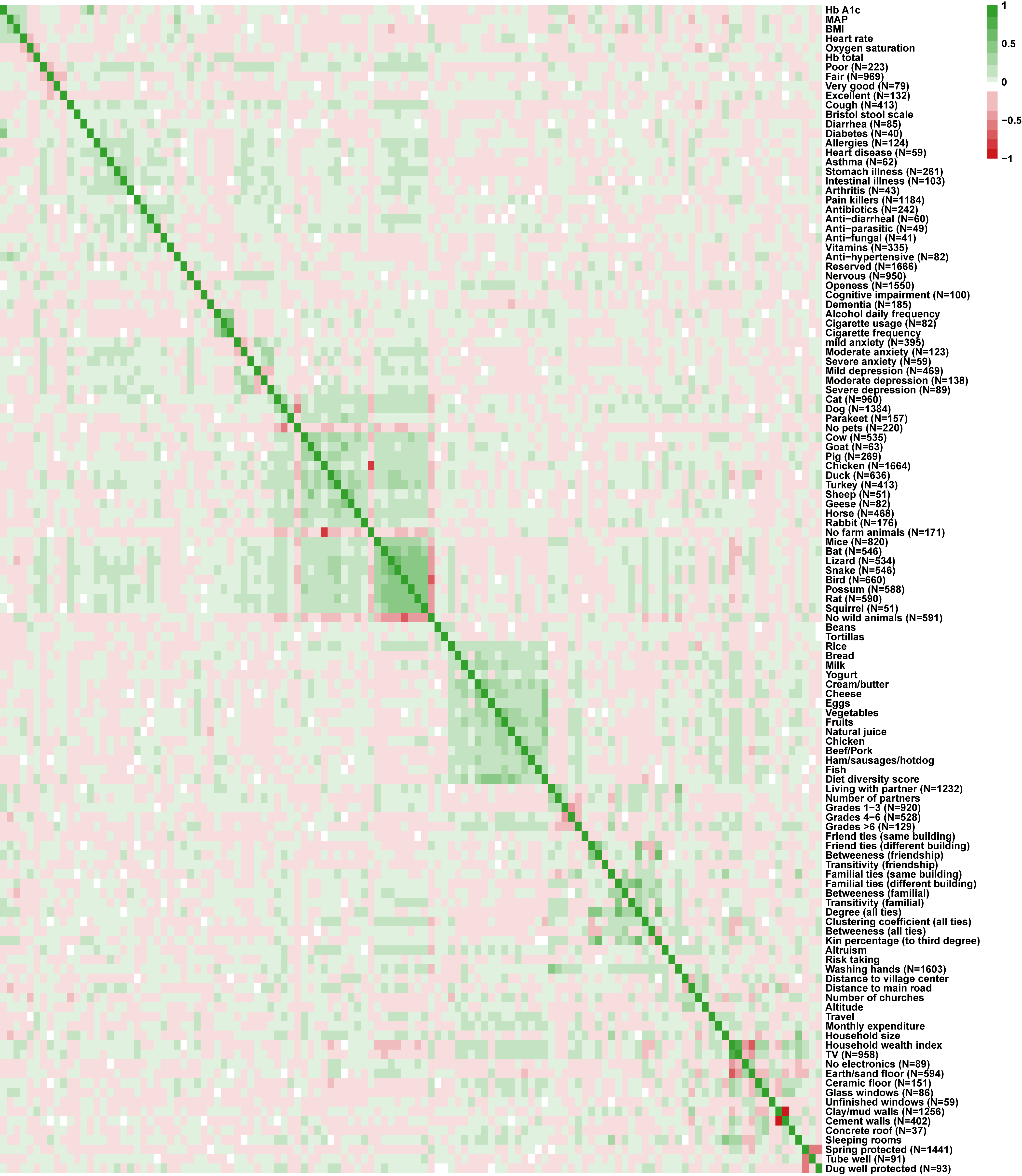

**Figure S1 Phenotype-phenotype correlation.** A matrix showing raw correlations between the phenotypes from every category (health, food and animals, socioeconomic factors). Column names are the same as the row names indicated on the right side of the matrix. Color ranges from positive (green) to negative (red) correlations.

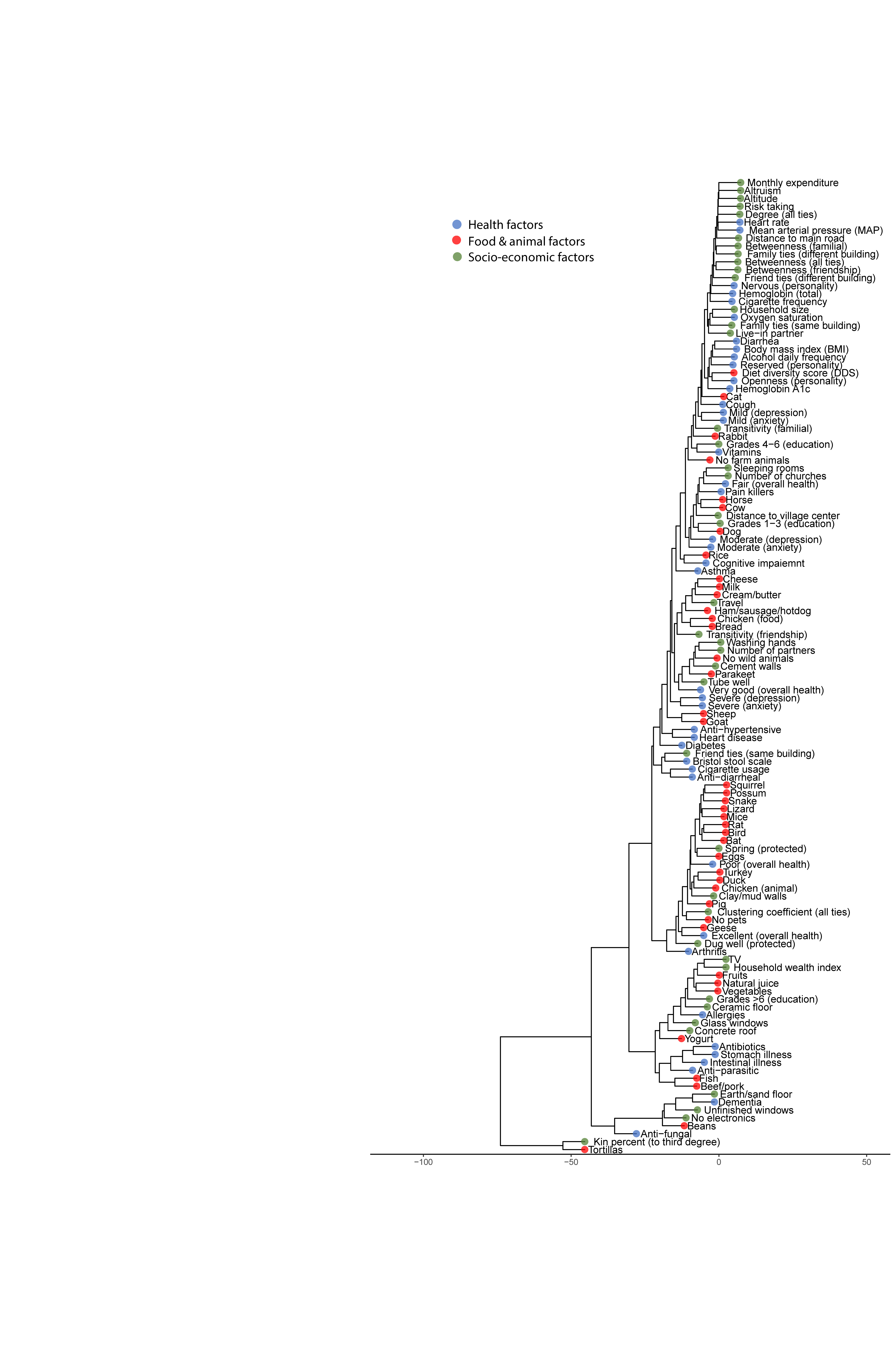

**Figure S2 Phenotype-microbiome association clustering.** Effect sizes from associations of all 123 phenotypes with 639 species are hierarchically clustered with respect to phenotypes. This phenotype tree is another representation of how similarly behaving a pair of phenotypes are with respect to how they associate with the gut microbiome overall.

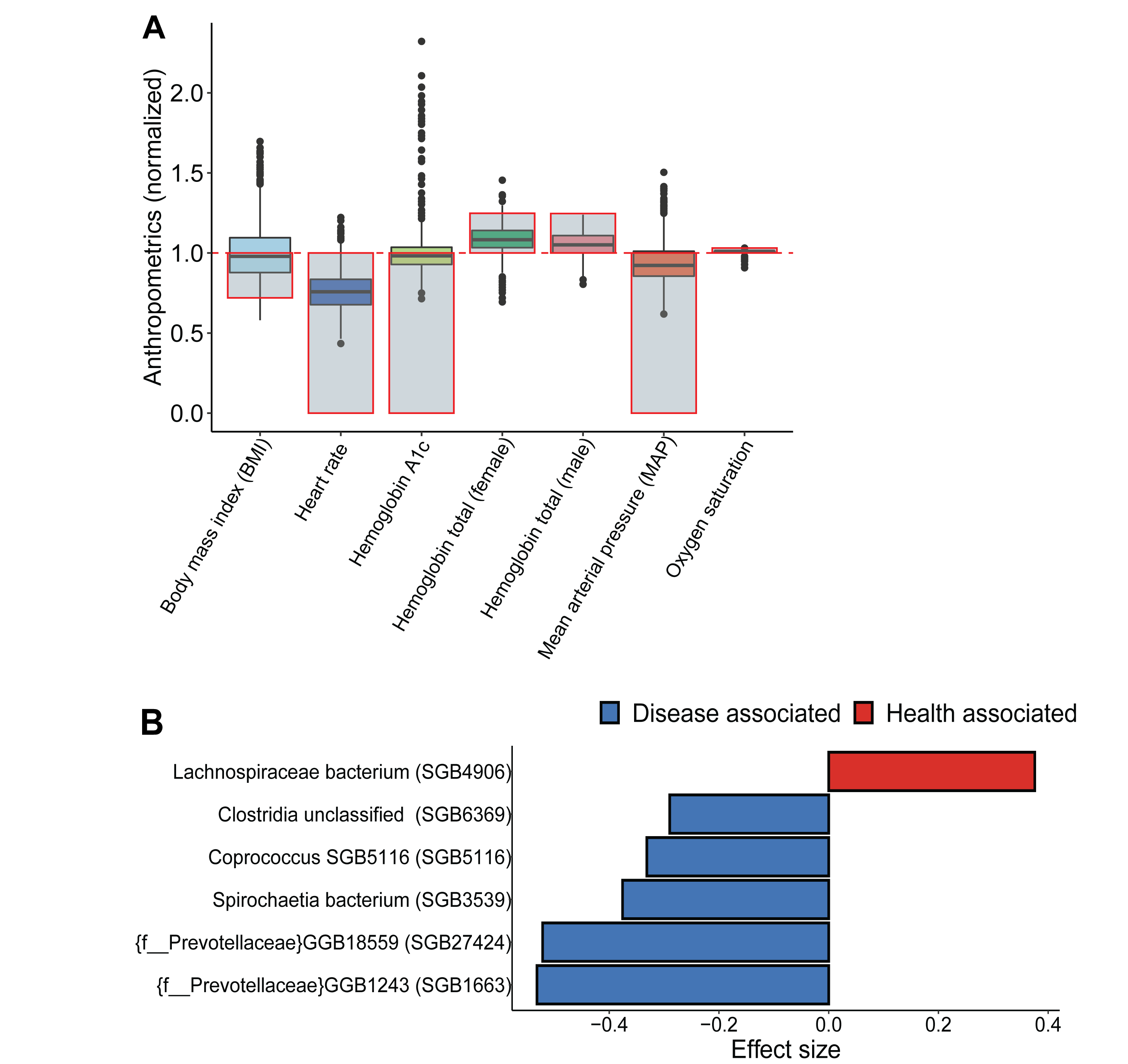

**Figure S3 Relationship between health and microbiome.** (A) Graphical visualization of physiological measurements (anthropometrics) of all N=1,871 villagers, with the grey box indicating normal values of each respective physiological measurement. The red box indicates the bounding limit of healthy ranges. (B) In the entire cohort, there were 468 chronically diseased individuals (who had at least one chronic condition). Differential abundance in healthy vs chronically diseased individuals using MaAsLin2 (see **Methods**) shows six significant species (after FDR correction of p-values). One of them (*Lacnospiraceae bacterium*) was differentially abundant in healthy individuals. On the other hand, five species (uSGB1663 and uSGB27424 from the *Prevotellaceae* family, *Spirochaetia bacterium*, *Coprococcus*, and uSGB6369 from the *Clostridia* family), two of which are unknown, were differentially abundant in chronically diseased individuals.

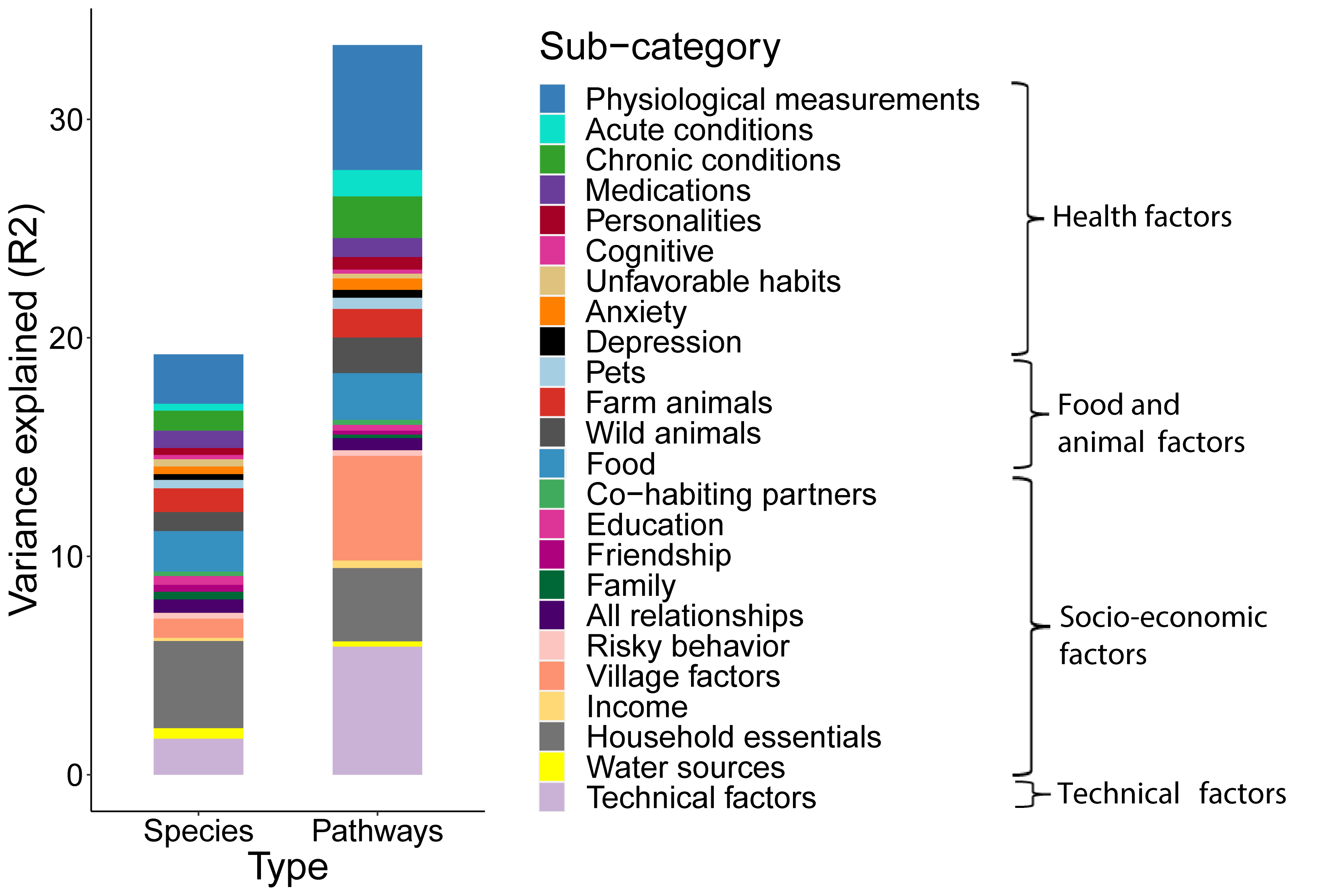

**Figure S4 Variance explained.** PERMANOVA analysis (999 permutations, p-value<0.001) computed on all phenotypes shows the variance explained in species and pathway compositions with a breakdown of sub-categories of all phenotypes (health, food and animal, socioeconomic factors). Overall, all the phenotypes together explain 19.2% and 33.4% of the variance explained in species and pathways, respectively. “Technical factors” here include age, sex, DNA concentration, sequencing batch, and sampling date. (See **Supplementary Table 7** for complete breakdown of variance explained in each sub-categories)

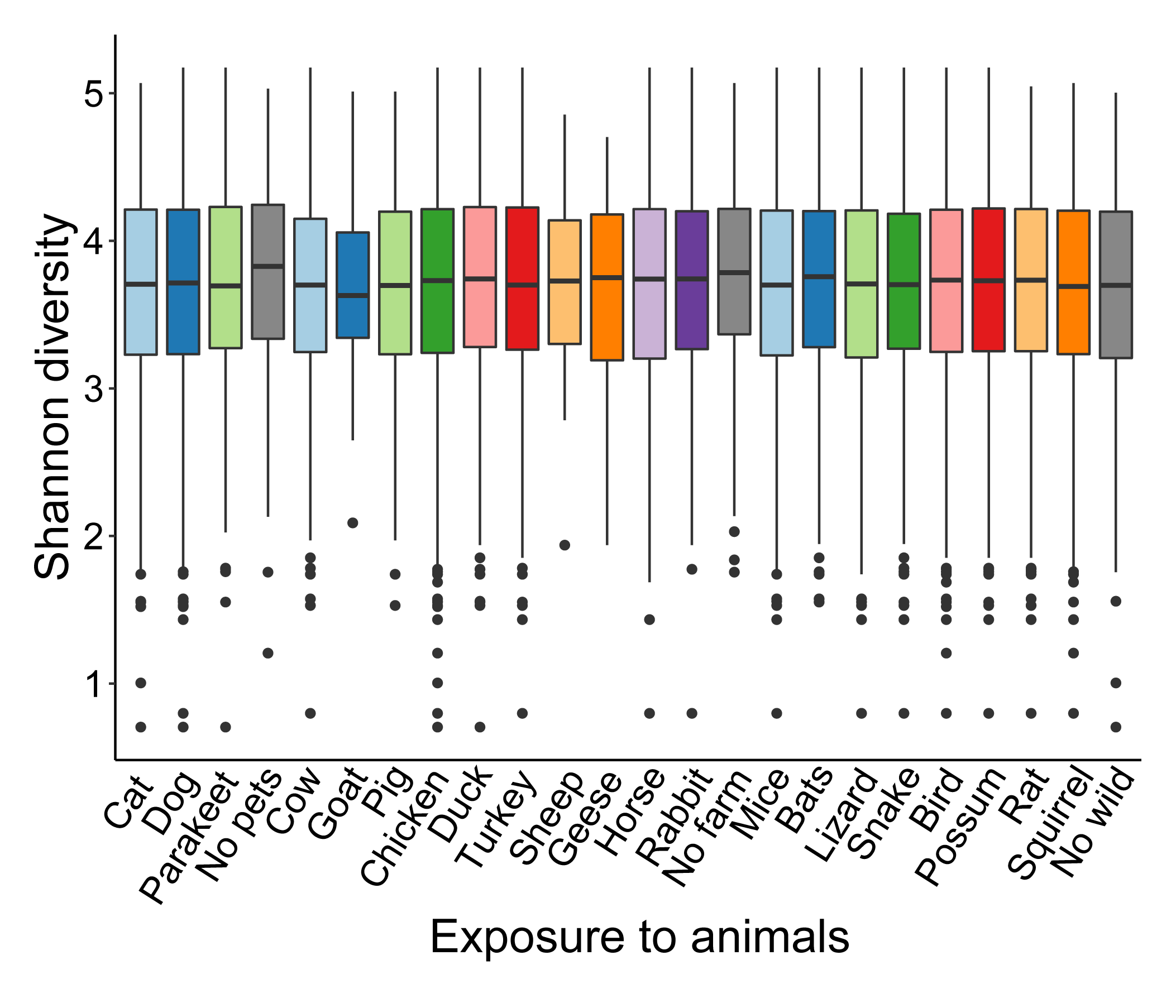

**Figure S5:** **Alpha diversity of individuals exposed to animals.**  Shannon diversity distribution among villagers exposed to pets, farm animals, and wild animals shows no significant differences between groups.

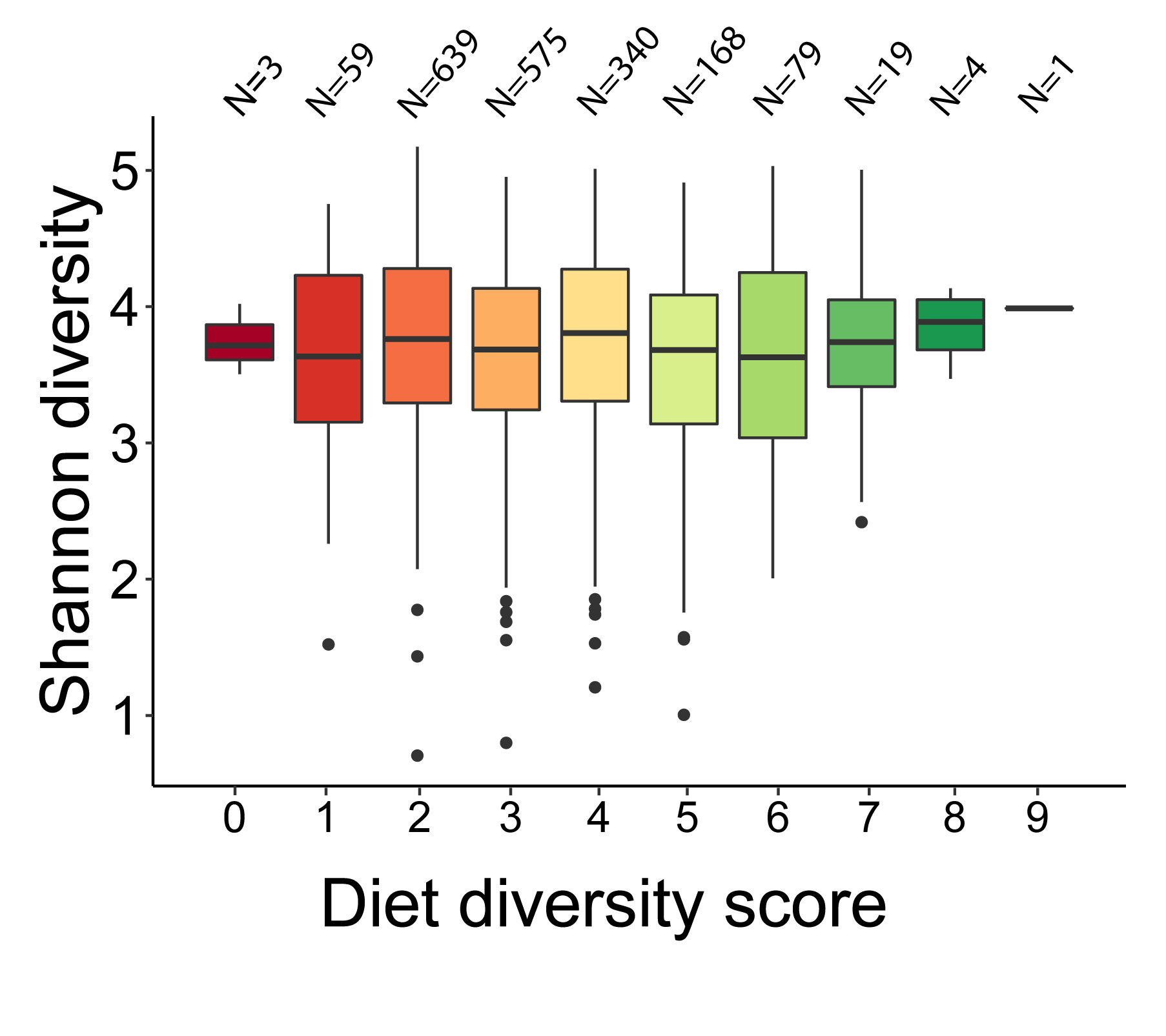

**Figure S6** **Diet diversity score.** Plot showing the Shannon diversities of individuals with varying diet diversity scores (see **Methods** for calculation of DDS scores).

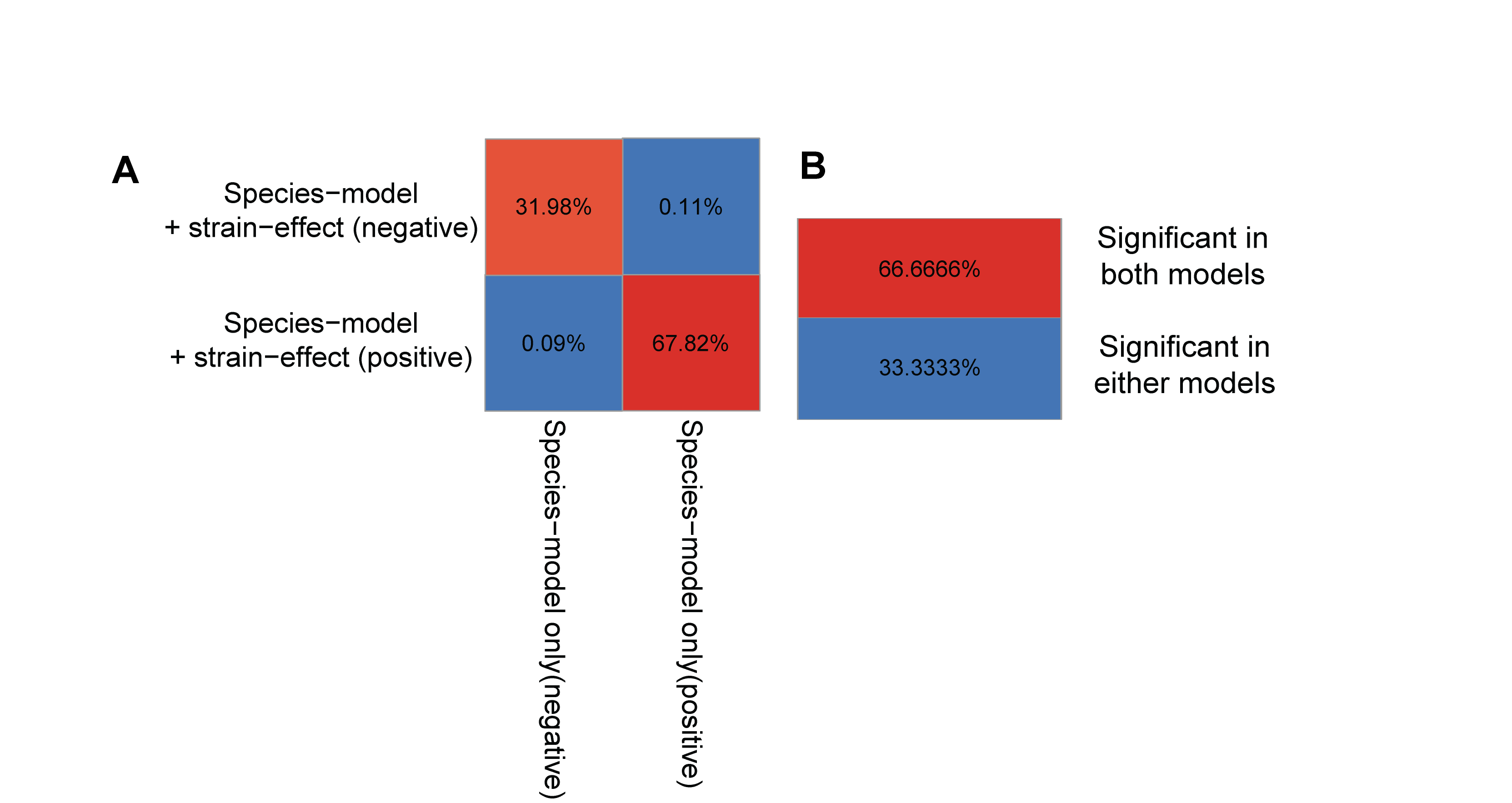

**Figure S7:** **Comparison of species and strain models.** (A) Side-by-side comparison of the direction of associations in both models (with and without strain-phylogenies). Each quadrant indicates positive or negative associations in either model. (B) Figure showing the presence of significant associations in both models compared to their presence in either of the models.

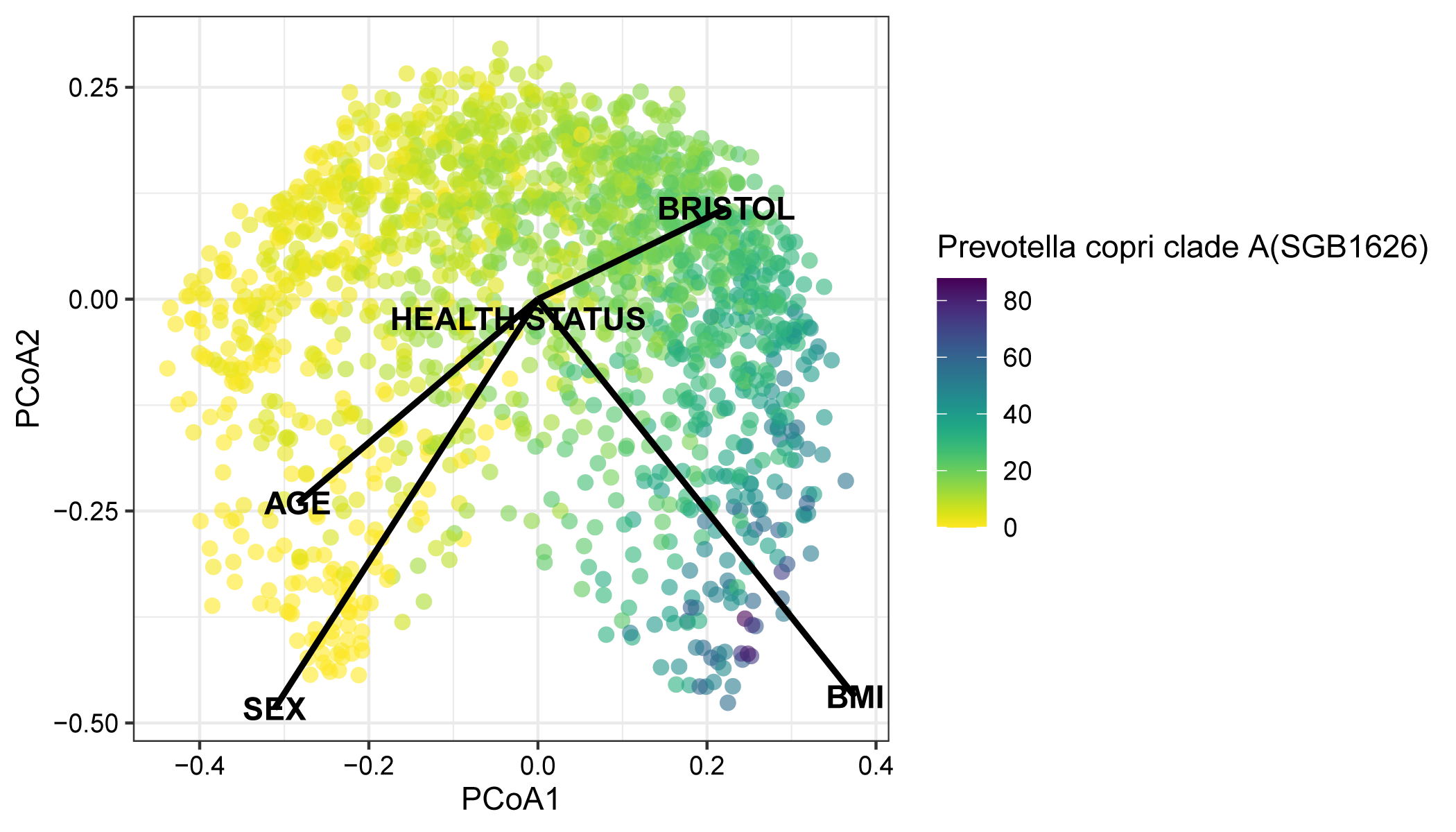

**Figure S8:** **Principal Coordinates Analysis (PCoA).** PCoA plot of the overall gut microbiome computed across 1,871 samples using the species-level relative abundances (legend) generated by MetaPhlAn4. Health status, age, sex, body mass index (BMI), and Bristol stool scale are shown as arrows along with the direction of influence. Samples are colored with the relative abundances of *Prevotella copri* (clade A).

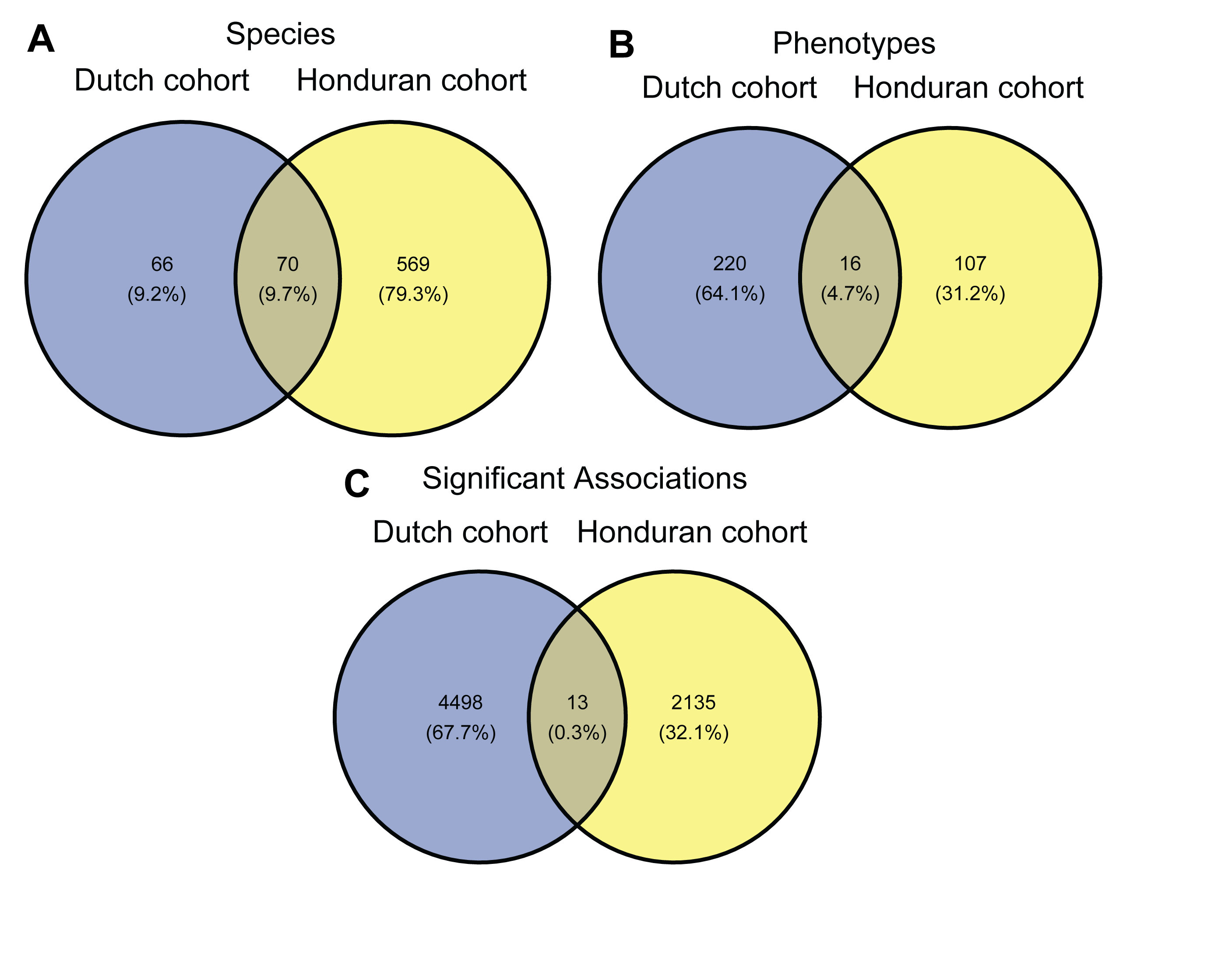

**Figure S9: Comparing Honduran and Dutch datasets.** We compared our findings to the Dutch Microbiome Project (LifeLines) dataset and we identified 13 associations in common. This relatively small number probably partially relates to differences in phenotypes measurement between the cohorts, and not just to the differences in the people and environments involved. Only certain phenotypes (N=16) were in common due to differences in phenotypic measurements and species profiling. For example, in the Dutch cohort, many measures captured variables like Arrythmia, Atherosclerosis, Heart failure, Heart rate complaints, and clogged arteries; but these were only measured as “Heart condition” in our Honduran dataset, which could include one or more of the foregoing variables. Similarly, in our Honduran dataset, we collect wealth variables in detail (e.g., owning a TV, refrigerator, metal or glass windows, number of sleeping rooms, and types of gas stoves), which could be reflected in variables like high/low-income proportion, or urbanicity in the Dutch dataset. In addition, the number of common species (N=70) was also a limiting factor in comparing across cohorts. All of these factors highlight the distinct dissimilarity in the two datasets (see **Supplementary Table 8** for a complete list of possible comparisons). (A) Honduran and Dutch datasets were profiled with different versions of MetaPhlAn (MetaPhlAn2 vs MetaPhlAn 4), resulting in newer species including uncharacterized species being identified in Honduran dataset (see **Methods**). (B)Less than 5% of the phenotypes were common to both datasets due to differences in measurement. (C) The above differences contributed to only 13 significant associations being common to both datasets.

**Supplementary Tables list:**

| **Supplementary Table** | | **Description** |
| --- | --- | --- |
| Supplementary Table 1 | Summary of associations between species and phenotypes | |
| Supplementary Table 2 | Summary of associations between pathways and phenotypes | |
| Supplementary Table 3 | Metadata summary of health phenotypes | |
| Supplementary Table 4 | Metadata summary of food & animal phenotypes | |
| Supplementary Table 5 | Metadata summary of socio-economic phenotypes | |
| Supplementary Table 6 | Summary of associations between species and unhealthy phenotypes | |
| Supplementary Table 7 | Variance explained in the gut microbiome | |
| Supplementary Table 8 | Comparison between Honduran and Dutch datasets | |
| Supplementary Table 9 | Summary of associations between species and phenotypes (along with strains) | |
| Supplementary Table 10 | Summary of associations between polymorphic sites and phenotypes | |

**Supplementary Table 1:**

Multivariate mixed linear regression between CLR transformed species (from MetaPhlAn 4) abundances and host-phenotypes of interest. (see **Methods**). Age, sex, BMI, Bristol stool scale, sampling date, batch effect, and DNA concentration are the covariates/fixed effects, and village is a random effect in the model. Effect sizes, p-values, and FDR-adjusted p-values for each pair of species-phenotypes are provided. In total 78,597 separate regressions were performed (639 species times 123 phenotypes).

**Supplementary Table 2:**

Multivariate mixed linear regression between CLR transformed pathway abundances (from HUMAnN 3) and host-phenotypes of interest. Age, sex, BMI, Bristol stool scale, sampling date, batch effect, and DNA concentration are the covariates/fixed effects and village is a random effect in the model. Effect sizes, p-values, and FDR-adjusted p-values for each pair of species-phenotypes are provided. In total 60,270 separate regressions were performed (490 pathways times 123 phenotypes).

**Supplementary Table 3:**

Summary description of health phenotypes stratified by sex and subcategories for all 1,871 individuals. (reported as percentages or mean (SD)). Subcategories include Physiological variables, Overall health, Acute conditions, Chronic conditions, Medication use, Personality types, Cognitive variables, Unfavorable habits, Anxiety, and Depression. Personality types have category types (agree strongly, agree, neither agree nor disagree, disagree, disagree strongly) (see **Methods**). Anxiety and Depression have mild, moderate, and severe ranges in their respective scales (GAD7 and PHQ9, see **Methods**).

**Supplementary Table 4:**

Summary description of food and animal phenotypes stratified by sex and subcategories for all 1,871 individuals. (reported as percentages or mean (SD)). Animal exposures are reported by percentage for all 3 sub-categories (Pets, Farm animals, and Wild animals). Individual food items are reported from FFQ (Food Frequency Questionnaire, see **Methods**).

**Supplementary Table 5:**

Summary description of socio-economic phenotypes stratified by sex and subcategories for all 1,871 individuals. (reported as % or mean (SD)). Social phenotypes include sub-categories like Partners, Education, and social-network factors (kin & non-kin).

Education has three levels: (Primary (1 - 3), Middle (4 - 6), and Secondary (>6). Environmental factors include risky behavior and village factors. Economic factors include Income, Household essentials, and Water sources. The Household wealth index (HHW) is constructed based on household essentials.

**Supplementary Table 6:**

Multivariate mixed linear regression between CLR transformed species abundances (from MetaPhlAn 4) and unhealthy ranges of health phenotypes of interest. All Unhealthy ranges were considered as separate levels in each regression, with healthy being the reference level. Age, sex, BMI, Bristol stool scale, sampling date, batch effect, and DNA concentration are the covariates/fixed effects, and Village is a random effect in the model. Effect sizes, p-values, and FDR-adjusted p-values for each pair of species-phenotypes are provided. In total 78,597 separate regressions were performed (639 species times 123 phenotypes).

Unhealthy ranges in Hemoglobin A1c can be categorized as: Diabetic (>7, 6.5 - 7) and Pre-diabetic (5.7 - 6.4). Unhealthy ranges in Blood pressure can be categorized as Elevated (Systolic (>119) & Diastolic (<=79), High blood pressure/Hypertension stage 1 (Systolic (>119) & Diastolic (>79)), and Hypertension Stage 2 (Diastolic (>89) or Systolic (>129)). Unhealthy ranges in Body Mass Index (BMI) can be categorized as Underweight (<18), Over-weight (25-30), Obese (30-35), and Morbidly Obese (>35). Unhealthy range in Heart rate is >100. Unhealthy ranges in Oxygen saturation include (<97). Unhealthy ranges in total Hemoglobin can be categorized as Elevated (male>18 or female>16) and Low (male<13 or female<12).

**Supplementary Table 7:**

Breakdown of the species and pathways variation explained by each sub-category belonging to health, food and animal, and socio-economic host phenotypes. Analysis was performed using PERMANOVA (999 permutations) (see **Methods**).

The total variance explained in gut microbiome species by host phenotypes is 26.39%, while the total variance explained in gut microbiome pathways by host phenotypes is 38.63%.

**Supplementary Table 8:**

Comparison of effect sizes, p-values, and FDR-adjusted p-values for each pair of species-phenotypes in the Honduran and the Dutch (LifeLines) datasets were closely examined for common associations. In total 13 significant associations were common between the two datasets. The disparity in the datasets is due to:
(i) Different numbers of species (due to different versions of MetaPhlAn used in profiling species abundances) led to 70 species being common to both datasets: and
(ii) Differences in measurements of host phenotypes across all categories led to 16 common phenotypes. (see **Figure S9**).

**Supplementary Table 9:**

Multivariate mixed linear regression between CLR transformed species abundances (from MetaPhlAn 4) and host-phenotypes of interest in samples having strain-phylogentic information (from StrainPhlAn). (see **Methods**). Strain phylogenies were constructed by StrainPhlAn for each species, resulting in a tree comprising of individuals based on the genetic makeup of the species in their respective guts. In addition to species-phenotype associations being performed (in samples present in the phylogenetic trees), the Almer package was used to add strain-phylogenetic information in the species-association model. Overall, species-phenotype association was contrasted with and without strains. Age, Sex, BMI, Bristol stool scale, sampling date, batch effect, and DNA concentration are the covariates/fixed effects, and village is a random effect in the model. Effect sizes, p-values, and FDR-adjusted p-values for each pair of species-phenotypes are provided. In total, 62,361 separate regressions were performed (507 species times 123 phenotypes). To avoid noise, StrainPhlAn trees with more than 100 samples was considered, resulting in 507 species.

**Supplementary Table 10:**

Multivariate mixed linear regression between polymorphic sites (from StrainPhlAn) and host-phenotypes of interest. (see **Methods**). Polymorphic sites were reconstructed by StrainPhlAn for each species, and its variation across samples was considered as a dependent variable in the regression model against host-phenotypes (independent variable). Overall, species-phenotype association was contrasted with and without strains. Effect sizes, p-values, and FDR-adjusted p-values for each pair of species-phenotypes are provided. In total 78,597 separate regressions were performed (639 species times 123 phenotypes).
